# Competitive Sports Participation in Athletes with Thoracic Aortic Aneurysms and Dissections

**DOI:** 10.1101/2025.10.14.25337926

**Authors:** Kayla House, Yasmin Toy, Nikhil Erabelli, Dipika Bhatia, Siddharth K. Prakash

**Affiliations:** John P. and Kathrine G. McGovern Medical School, University of Texas Health Science Center at Houston, Houston, TX, USA; Tulane University School of Medicine, New Orleans, LA, USA

**Author notes:** Corresponding author: Siddharth Prakash, MD, PhD, Department of Internal Medicine, The University of Texas Health Science Center at Houston, 6431 Fannin Street, MSB 6.116, Houston, Texas 77030. Kayla House, Tulane University School of Medicine, 1430 Tulane Avenue, New Orleans, Louisiana.

**Keywords:** thoracic aortic aneurysms and dissections, competitive sports, health outcomes

## Abstract

**Objectives:** Individuals with thoracic aortic disease (TAD) are frequently counseled to avoid intensive exercise. We surveyed athletes with TAD to assess their athletic participation and cardiovascular symptoms or outcomes related to competitive sporting events.

**Methods:** Participants were eligible for this study if they had a diagnosis of TAD and had participated in at least one competitive athletic event. Data was collected via customized REDCap surveys. The primary study outcomes were competitive event participation and event-related symptoms, comparing the five years before and after aortic diagnosis.

**Results:** Anonymous surveys were sent to a total of 8,300 participants in two social media groups. Eighty-six eligible participants completed surveys: 37 with aneurysms, 33 with dissections, and 16 with both diagnoses. After diagnosis, competitive sports participation decreased from a mean of 7 events per year (10% of cohort) to a mean of 0.8 events per year (1.2% of cohort, P<0.05). Seven respondents experienced event-related health symptoms, primarily related to running or endurance events. One participant was hospitalized after competition for arrhythmia. No acute dissections or aortic interventions occurred. Weekly exercise time was greater in those without event-related symptoms. In multivariate analyses, the number of cardiovascular medications was associated with event-related symptoms.

**Conclusion:** Athletic competition decreased significantly after TAD diagnosis. In a selected cohort of highly active individuals with TAD, competition did lead to frequent symptomatic episodes but did not result in acute aortic events. These observations reinforce the importance of regular exercise to prevent symptomatic cardiovascular events for TAD patients who persist in competition.

**Lay Summary:** This study explored how people with thoracic aortic disease (TAD) who regularly competed in sports changed their participation after diagnosis and whether athletic events triggered any heart- or vessel-related problems.

- Key Finding 1: Competitive sports participation dropped sharply after diagnosis.
- Key Finding 2: While some participants experienced symptoms such as shortness of breath or irregular heart rhythm during endurance events, no one suffered an acute aortic tear or required emergency aortic surgery.

## INTRODUCTION

Individuals who are diagnosed with thoracic aortic aneurysms (TAA) or aortic dissections (AD), collectively known as thoracic aortic disease (TAD), are generally counseled to avoid competitive and contact sports [1–3]. Competitive athletic activities are frequently associated with high intensity exertion and an expectation that participants will push themselves to their physical limits, but this concept does not apply to all sports [1]. For example, competition includes both low intensity (e.g. golf, bowling) and high intensity activities (running, singles tennis) [1]. Moreover, athletes are motivated to participate in competitive sports for different reasons, such as a desire for self-improvement or a drive to demonstrate superiority [4,5].

Competitive athletic activities can also be distinguished by the predominance of dynamic (isotonic) or static (isometric) components [1,6,7]. In isotonic exercises, such as bicep curls or pushups, muscles lengthen or shorten rhythmically, resulting in a relatively small intramuscular force and increased cardiac output. Isometric exercises, such as wall sits or planks, involve stationary muscle contractions and relatively large intramuscular force [8]. In addition, the Metabolic Equivalent of Task (MET) is a widely used classification for exercise intensity and is based on the energy required to perform physical activities [6,7]. Therefore, the cardiovascular effects of exercise are related to the intensity and type of athletic activities.

Routine exercise stimulates cardiac remodeling [1,8–11]. Myocardial cells respond to exercise by increasing contractile force and stroke volume, and over time repetitive training tends to increase ventricular efficiency during exercise. Studies of humans and animal models have documented increases in myocardial mass and ventricular dilation after prolonged exercise training that may overlap with cardiomyopathic values but appear to carry a benign prognosis [8–12]. For example, mild thoracic aortic dilation is frequently observed in competitive athletes but is usually below the threshold for diagnosis of an aneurysm [10]. In Marfan syndrome mouse models, aortic dilation rates were decreased to near wild-type levels in comparison to sedentary controls when mice were exposed to 60 minutes per day of moderate intensity aerobic exercise [9,13]. These findings suggest that aortic enlargement and other cardiac structural changes in response to exercise may be appropriately adaptive because they do not predict future cardiovascular events [1,11]. Moreover, increased physical conditioning due to exercise training decreases heart rate and systolic blood pressure, a major risk factor that when uncontrolled may increase the penetrance and severity of thoracic aortic disease [14].

In the absence of evidence-based guidelines, patients with TAD are frequently counseled to restrict their physical activities, including competitive sports participation [1,14]. Engaging in competition for self-improvement (e.g. as a benchmark to attain personal goals) can provide a powerful incentive to pursue physical activities. Therefore, decreased sports participation may negate motivation for all types of exercise.

Many individuals who are diagnosed with TAD are prescribed antihypertensive medications that may change their physiologic responses to exercise. Evidence evaluating the protective or detrimental effects of cardiovascular medications during competitive activities is urgently needed to manage individuals with a history of thoracic aortic disease who engage in competition [1,15].

## MATERIALS AND METHODS

### Ethics

The Committee for the Protection of Human Subjects at the University of Texas Health Science Center at Houston determined that this study meets the criteria for exemption research. Therefore, the regulatory requirements for informed consent do not apply.

### Study Design

This study is an observational cohort study. Associations between health outcomes and clinical or exercise variables were evaluated using t-tests, one-way ANOVA, and logistic regressions.

### Participants and Setting

Respondents were recruited from the Aortic Athletes (797 members) and the Cardiac Athletes (7500 members) Facebook groups. Individuals were eligible for the study if they had survived a thoracic aortic dissection, were diagnosed with a thoracic aortic aneurysm, and had participated in at least one competitive athletic event prior to their initial aortic event. Surveys were distributed anonymously using a customized version of REDCap at the University of Texas Health Science Center at Houston. Respondents were provided with a prompt outlining the purpose of the study and a definition of competitive events.

### Surveys

The primary REDCap survey consisted of 20 questions about eligibility, demographics, TAD characteristics, competitive sports participation before and after diagnosis, weekly estimates of exercise time, and event-related health outcomes (Supplemental Data).

A separate survey that was shared only with the Aortic Athletes group included 20 questions about exercise habits, attitudes toward exercise, mental health, and physician messaging about exercise (Supplemental Data). This survey specified the types of exercise performed by respondents and assessed both the amount of exercise and intensity level before and after dissection.

Exercise intensity was classified according to MET thresholds based on the 2024 Compendium of Physical Activities: <3 low, 3-6 moderate, >6 high [1,16].

### Data Analysis

Survey data was exported directly from REDCap into a secure spreadsheet. Survey responses were analyzed using descriptive statistics (frequencies, percentages, means, medians, standard deviations where appropriate). Student t-tests and one-way ANOVA tests were performed to evaluate associations between respondent characteristics and event-related health symptoms. One-way ANOVA and Pearson correlation tests were used to evaluate differences in competitive sports participation before and after the diagnosis of TAA or TAD. All quantitative analyses were conducted in Microsoft Excel (v16.85).

### Data Availability

All data generated or analyzed during this study are included in this published article and supplementary information files.

## RESULTS

### Attitudes Toward Exercise

A preliminary survey was distributed only to the Aortic Athletes Facebook group with 114 responses. 72% of respondents reported that they experienced significant anxiety or frustration about exercise after diagnosis. More than 80% of respondents had received directives from their physicians about limiting activities such as weightlifting (*N* = 69), Valsalva maneuvers (*N* = 24), contact sports (*N* = 7), or high-intensity exercises (*N* = 46). The most frequent concerns of respondents were that specific exercise restrictions were inconsistent between providers or were not clearly communicated.

### Study Cohort

The main survey about competitive sports participation was distributed to the Aortic Athletes and Cardiac Athletes Facebook groups. A total of 86 surveys from both groups were eligible for analysis (Table 1). The primary diagnoses of survey respondents included thoracic aortic dissection (*N* = 33), thoracic aortic aneurysm (*N* = 37), and both diagnoses (*N* = 16). Running was the most common competitive event (Table 2). Respondents participated in fewer competitive sporting events after TAD (Figure 1).

**Table 1.** Characteristics of Survey Respondents. TAD: thoracic aortic dissection; TAA: thoracic aortic aneurysm; Proximal: location of TAA in sinuses of Valsalva and/or ascending aorta; CV: cardiovascular. Values are *N* (%), mean (standard deviation).

| Variable | N (% or mean) |
| --- | --- |
| **Age (years)** | **54 (28)** |
| **Age at TAD diagnosis** | **48 (11)** |
| **TAD** | **33 (38)** |
| Type A | 11 (52) |
| Type B | 2 (10) |
| More than one dissection | 8 (38) |
| **TAA** | **37 (43)** |
| Proximal | 31 (91) |
| Abdomen | 3 (8) |
| Other Location | 12 (33) |
| **TAA and TAD** | **16 (19)** |
| **Years After Diagnosis** | **4.2 (4.8)** |
| Years after TAD | 5.1 (5) |
| Years after diagnosis of TAA | 2.8 (2) |
| **Aortic diameter (centimeters)** | **5.6 (5.2)** |
| **Number CV medications** | **1.3 (0.8)** |

**Table 2.** Comparison of Study Participants With and Without Event-Related Symptoms. Values are presented as means (standard deviations). Min: minutes; Wk: week; cm: centimeters; CV: cardiovascular.

| Variable | Symptoms<br>(N = 7) | No Symptoms<br>(N = 79) | *P* |
| --- | --- | --- | --- |
| Training time (min/wk) | 222.1 | 430.6 | 0.10 |
| Recreational time (min/wk) | 120.0 | 276.3 | 0.10 |
| Total exercise time (min/wk) | 273.6 | 552.6 | 0.03 |
| Frequency of training (days/wk) | 4.75 | 4.84 | 0.48 |
| Length of training sessions (min) | 73.2 | 110.5 | 0.15 |
| Age at dissection (years) | 42.6 | 48.8 | 0.06 |
| Aneurysm size (cm) | 4.75 | 4.80 | 0.45 |
| Number of CV medications | 2.29 | 1.19 | 0.01 |

**Figure 1.**
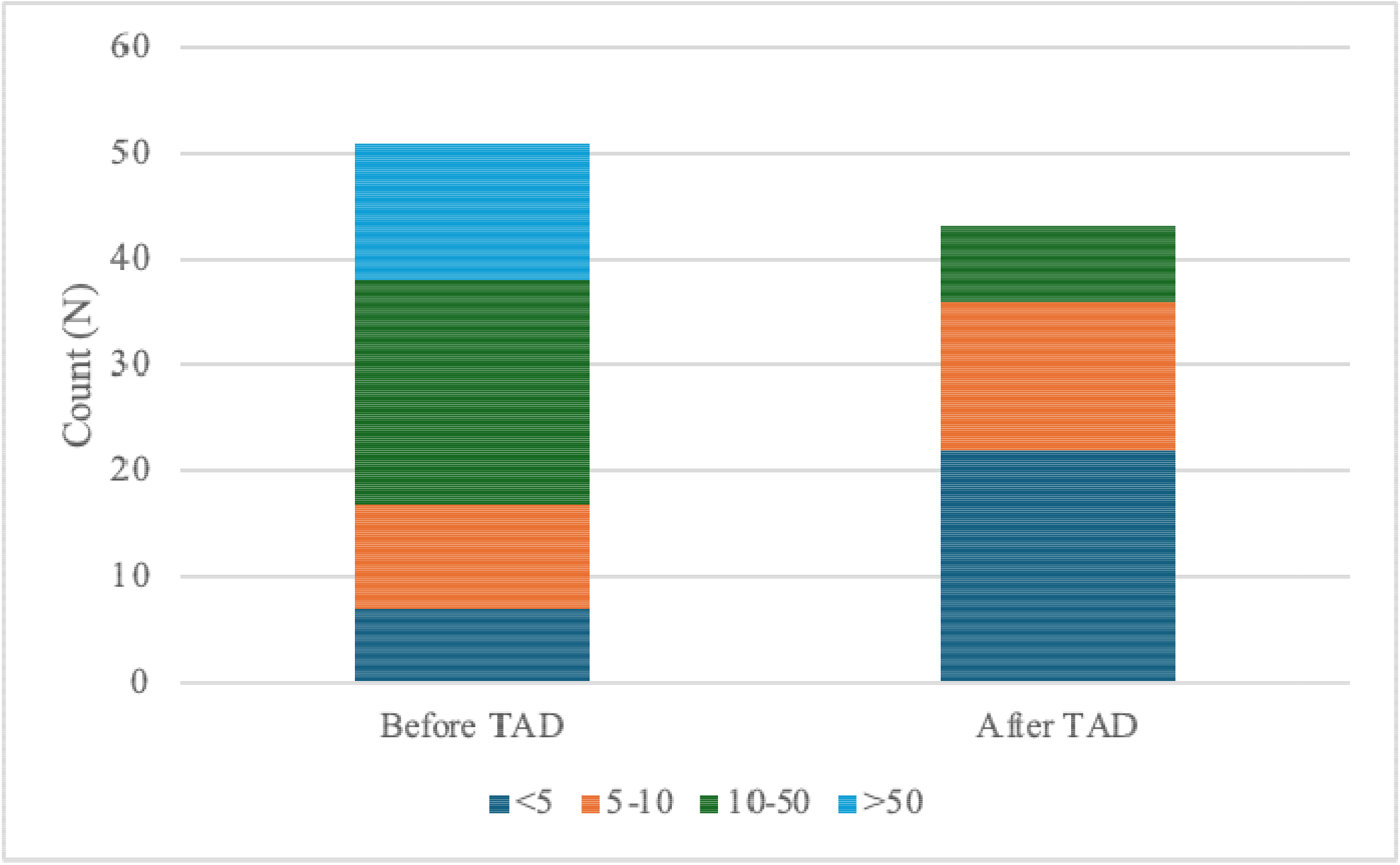
Total number of competitive sports events in the five years before and five years after TAD diagnosis.

### Event-related symptoms

Seven participants (10%) experienced event-related health symptoms. Event-related health symptoms included syncope or presyncope (*N* = 10), visual disturbance (*N* = 2), arrhythmia (*N* = 2), chest pain (*N* = 1), and one hospitalization (Supplemental Table 2). Respondents who experienced event-related health symptoms spent an average of 295 minutes per week on combined training and recreational exercise time and took an average of 2 cardiovascular medications (Table 2). In logistic regression analysis, the number of cardiovascular medications was the only significant predictor of event-related symptoms (OR 9.69, 95% CI: 1.10, 91.7). Exercise time was not an independent predictor of event-related symptoms as a continuous variable (OR 2.9, 95% CI: 0.53-16.03) or as a dichotomous variable (AUC = 0.693).

## DISCUSSION

The safety and health outcomes of athletic competition for patients with TAD is largely unknown because they are often counseled to avoid vigorous exercise. To understand the extent of athletic participation after the diagnosis of TAD, we surveyed 107 individuals with TAD who had competed in at least one lifetime competitive sporting event. The objective of this study was to assess the range and intensity of competitive sports involvement and related health symptoms in athletes with TAD. The mean age of the study cohort was 54 years, more than 10 years younger than the IRAD registry cohort, but generally representative of TAD patients, who are most frequently ascertained between ages 50 and 60 [17]. We found that competitive sports participation decreased significantly in the five years following the diagnosis of TAD. This trend reflects widespread counseling by physicians to avoid vigorous exercise and anxieties about exercise that were reported by many participants. In compliance with current guidelines, most athletes avoided isometric events and competed in intense aerobic sports such as running, cycling, basketball, and endurance events. Overall, we found that supervised moderate intensity exercises were not associated with new aortic dissections or surgical interventions. Only seven of 86 respondents reported event-related health symptoms, and individuals who did not report event-related symptoms (N = 79) reported significantly more regular weekly exercise time than those who reported event-related symptoms (*p* = 0.03). Thus, we suggest that relative deconditioning was the most probable cause of event-related symptoms. While cardiovascular medications were associated with symptomatic episodes, additional studies are needed to determine how cardiovascular fitness, medications, and thoracic aortic disease may interact to cause exertional symptoms.

### Limitations

We did not collect data about underlying heritable thoracic aortic diseases (HTAD) such as Marfan syndrome or Loeys-Dietz syndrome. It is unclear if the long-term outcomes of exercise are different in individuals with heritable thoracic aortic diseases compared to participants who do not have a genetic diagnosis, and it is possible that a disproportionate number of respondents with event-related symptoms also had HTAD. There were insufficient numbers of participants who engaged in non-aerobic competitive activities to reach conclusions about their safety.

Surveys did not collect data about biological sex or gender, which could potentially impact the risk for event-related symptoms. This self-reported data from a highly selected group of participants may be subject to substantial bias and should not be generalized to all individuals with thoracic aortic disease without validation in additional studies.

### Conclusion

In a highly active cohort of individuals with TAD, repetitive participation in competitive athletics did not lead to dissections or surgical interventions but was associated with relatively frequent adverse health events and at least one hospitalization. However, there were no reported dissections or surgical interventions. Overall competitive sports participation declined after diagnosis. We also observed that the number of cardiovascular medications and the intensity of the static and dynamic components of sports events may contribute to event-related symptoms. The data suggests that increased physical conditioning may be beneficial to prevent event-related health symptoms, especially in high-intensity competitive sport participants. Therefore, counseling about proper conditioning prior to competition, in addition to careful review of cardiovascular prescriptions, may help to reduce event-related health symptoms. Additional research is needed to determine the optimal medication and training regimen for safer outcomes in individuals with TAD who are active in competitive sports.

## ACKNOWLEDGEMENTS

We are profoundly grateful to Jack Crowe, Carmen David, the Aortic Athletes, and the Cardiac Athletes, the Genetic Aortic Disorders Association (GADA) Canda, and the John Ritter Foundation for their continual support of our work. The results of our study are presented clearly, honestly, and without fabrication, falsification, or inappropriate data manipulation. The results of our study do not constitute endorsement by the American College of Sports Medicine.

## AUTHOR CONTRIBUTIONS

Dipika Bhatia: Data analysis, Writing – Reviewing and Editing; Nikhil Erabelli: Data analysis, Writing – Reviewing and Editing; Kayla House: Data curation, Writing – Original draft preparation; Yasmin Toy: Writing – Reviewing and Editing; Siddharth Prakash: Conceptualization, Methodology, Writing – Reviewing and Editing, Project administration.

## LIST OF SUPPLEMENTAL DIGITAL CONTENT

Table 1: Erabelli.Bhatia.Prakash.MSSE.Table1 (Word document)

Table 2: Erabelli.Bhatia.Prakash.MSSE.Table2 (Word document)

Figure 1. : Erabelli.Bhatia.Prakash.MSSE.Figure1 (Word document)

Supplemental Material: Erabelli.Bhatia.Prakash.Supplement (PDF)

