## Supplemental Data for "Competitive Sports Participation in Athletes with Thoracic Aortic Aneurysms and Dissections"

Aortic Athletes Survey

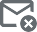

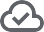

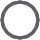

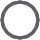

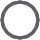

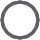

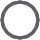

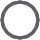

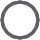

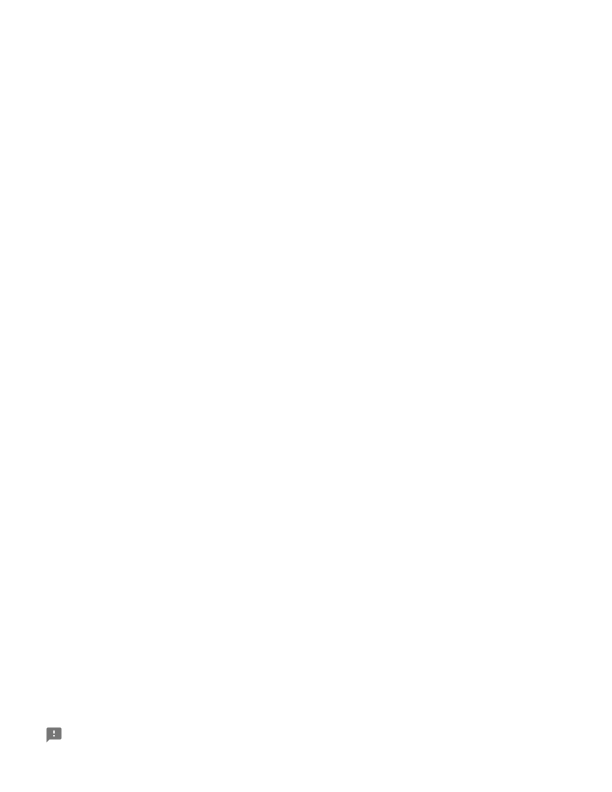

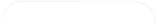

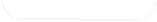

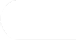

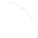

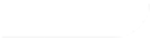

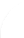

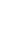

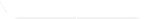

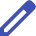

1 of 6

This is an anonymous survey to collect feedback on the Aortic Athletes group. Thank you for your time!

Not shared

* Indicates required question

### What is your diagnosis of aortic disease?

Post surgical repair of aneurysm Dissection of aorta

Dilation or aneurysm (watch and wait)

Other:

### If you have experienced an aortic dissection, was it:

Repaired through surgery (either open heart or endovascuar) Medically managed/no surgery

Both surgically repaired and a remaining medically managed aneurysm

### What was your age at diagnosis?

Your answer

### What is your current age?

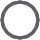

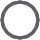

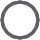

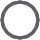

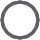

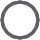

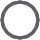

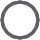

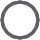

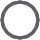

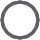

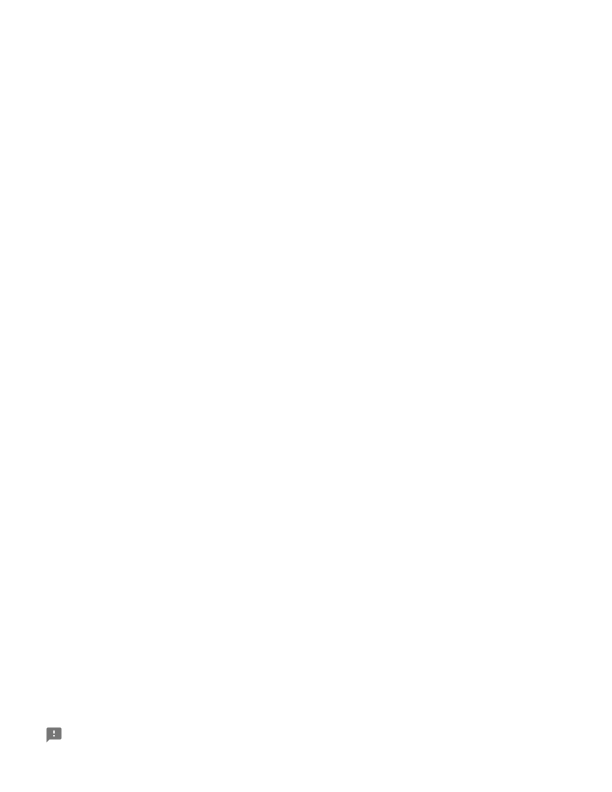

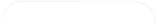

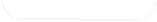

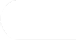

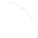

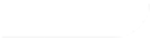

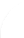

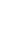

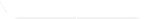

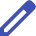

Your answer

### What is your country of residence?

Your answer

### What activity serves as your primary exercise?

Walking Running Biking Hiking Weightlifting Swimming Golf

Other:

### Approximately how long have you been a member of Aortic Athletes (which started in March 2023)?

More than 1 year

Between 6 months and 1 year Less than 6 months

2 of 6

### Since joining Aortic Athletes do you:

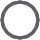

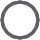

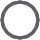

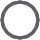

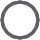

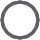

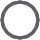

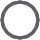

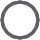

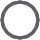

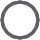

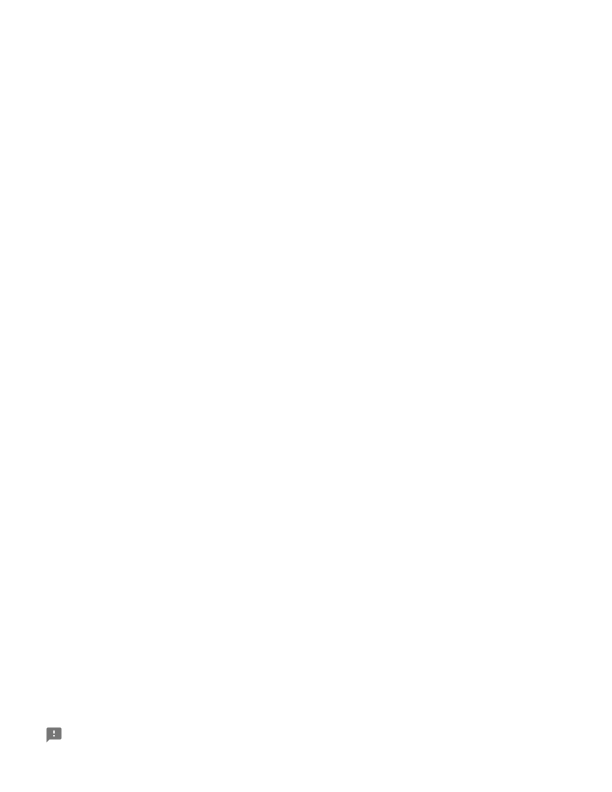

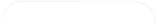

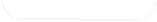

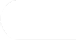

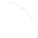

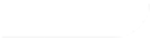

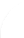

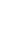

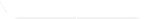

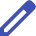

3 of 6

Exercise less Exercise more

Exercise approximately the same amount as before joining

### BEFORE being diagnosed with aortic disease, approximately how many minutes per week did you exercise?

None/Zero

Some, but less than 150 minutes per week 150 minutes to 300 minutes per week More than 300 minutes per week

### AFTER joining Aortic Athletes, approximately how many minutes per week did you exercise?

None/Zero

Some, but less than 150 minutes per week 150 minutes to 300 minutes per week More than 300 minutes per week

### BEFORE joining Aortic Athletes, what was your typical intensity level during exercise?

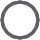

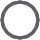

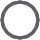

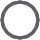

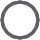

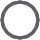

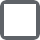

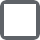

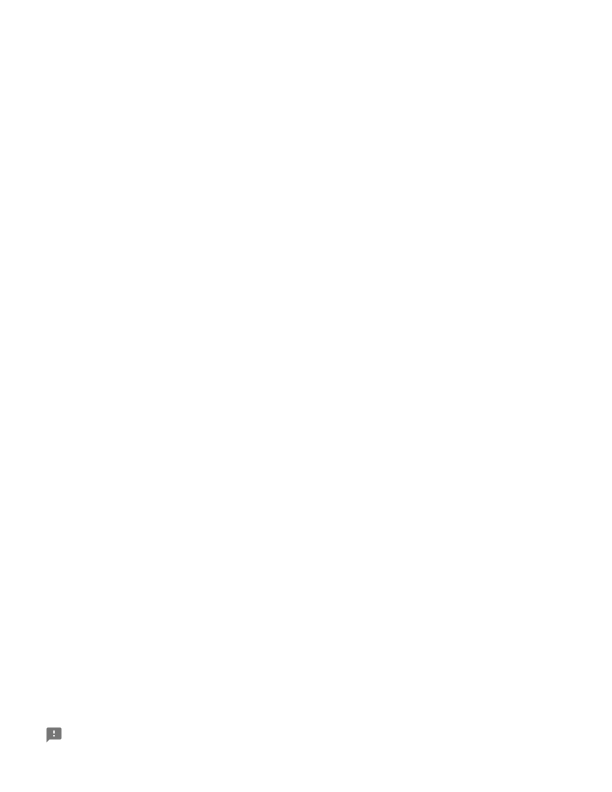

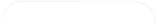

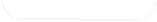

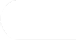

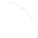

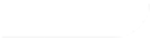

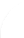

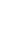

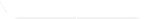

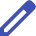

4 of 6

Light Moderate Vigorous

### AFTER joining Aortic Athletes, what is your typical intensity level during exercise?

Light Moderate Vigorous

### What type of exercise do you engage in?

Aerobic (examples: walking, running, swimming, biking)

Strength training (examples: lifting weights, body weight exercises including yoga and Pilates)

### Have your physician(s) given you limitations on exercise? If yes, brieﬂy describe (less than 100 words) those physician limitations

Your answer

### Did you experience any fear of exercise BEFORE joining Aortic Athletes based on your diagnosis of aortic disease or any limitations placed by your physician?

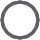

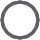

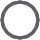

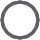

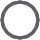

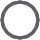

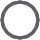

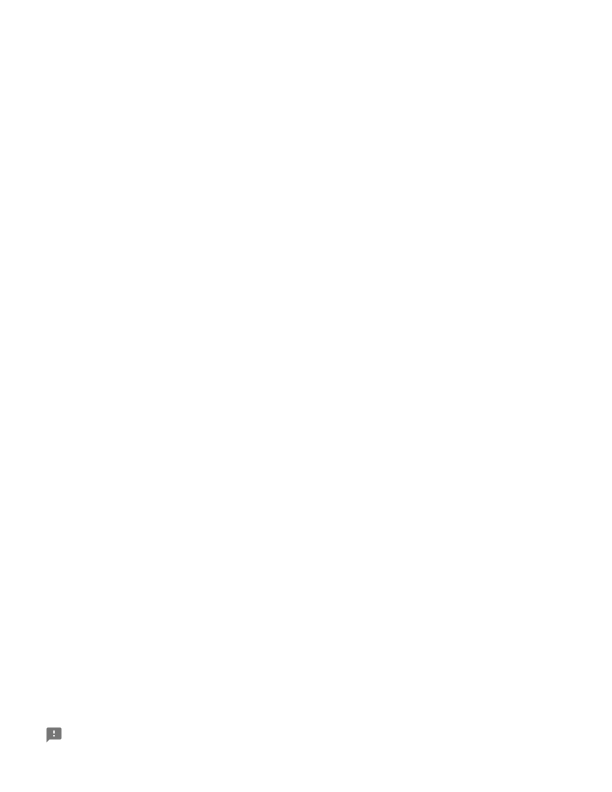

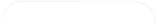

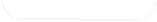

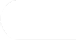

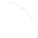

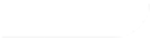

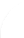

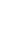

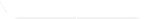

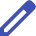

5 of 6

Yes No

### If you experienced a fear of exercising before joining Aortic Athletes, is that fear

More Less Same

### If you answered less fear since joining Aortic Athletes, what caused the change? (In 100 words or less)

Your answer

### Have you participated in an Aortic Athletes exercise challenge?

Yes No

### If yes, did the exercise challenge

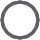

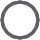

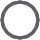

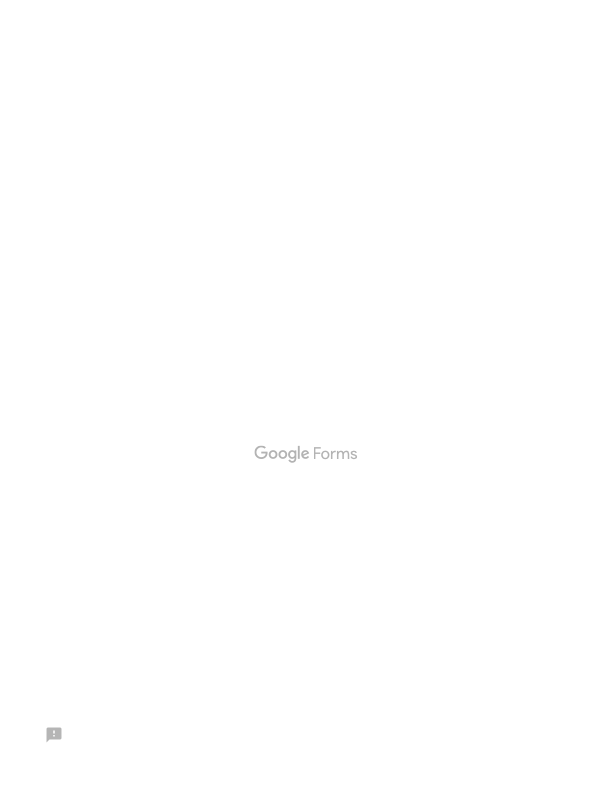

6 of 6

Increase the amount you exercise Decrease the amount you exercise

Your exercise volume remained the same

### If you have not participated in an Aortic Athletes exercise challenge, what are the reasons you have not participated? (Less than 100 words)

Your answer

Submit	Clear form

**Thoracic Aortic Dise ase: C ompet itive At hletic Even ts After Diagnosis**

*Page 1*

The purpose of this study is to collect data from individuals who have:

- been diagnosed with aortic aneurysm OR survived an aortic dissection
- participated in competitive athletic events

Note: competitive athletic events include ALL officially timed events such as marathons, 5K's, etc. Please read each question carefully before answering.

Thank you!

-Dr. Siddharth Prakash and Team at UTHealth Houston

I have an:	Aortic aneurysm

Aortic dissection Both

None of the above

How many years ago was your dissection?

(Example: 5 years)

How old were you when your dissection occurred?

**Please read the following definitions before answering the next question:**

Type A dissection: affects the part of the aorta closest to the heart (top of the candy cane)

Type B dissection: affects the part of the aorta further from the heart (bottom of the candy cane)

What type of dissection did you have?	Type A Type B

Type A and B I don't know

□	projectredcap.org

*Page 2*

How many years ago were you diagnosed with an aortic aneurysm?

Where is your aneurysm?	Aortic root (beginning part of aorta) Somewhere else in my chest Abdomen

I don't know

How large is your aneurysm in centimeters? If you don't know, leave this blank.

Have you ever participated in a competitive sports	Yes

event (in your entire life)?	No

Examples: marathon or any sports event that is timed, tennis tournament, organized sports game of any kind

Approximately how many competitive sports events have	0

you ever participated in?	Less than 5

5-10

10-50

More than 50

Thinking about the 5 years before your dissection or	0

diagnosis of an aneurysm, how many competitive	Less than 5

sporting events did you participate in?	5-10

10-50

More than 50

Note: If you had both a dissection and an aneurysm, count the time from the one that happened first.

Thinking about the 5 years after your dissection or	0

diagnosis of an aneurysm, how many competitive	Less than 5

sporting events did you participate in?	5-10

10-50

More than 50

Note: If you had both a dissection and an aneurysm, count the time from the one that happened first.

**The next questions only apply to competitive events that you participated in after your**

**dissection or diagnosis of an aneurysm:**

Which competitive events did you participate in?	Running

Check all that apply.	Ironman or similar endurance event Cycling

Examples: marathon or any athletic event that is	Basketball timed, tennis tournament, organized sports game of any	Football kind	Soccer

Tennis Dancing

Swimming

Other (please specify):

□	projectredcap.org

*Page 3*

Which of the following types of cardiovascular	Beta blocker (metoprolol, atenolol, carvedilol) medications were you taking when you participated in	Angiotensin receptor blocker (losartan, valsartan, previous sporting events? Check all that apply:	irbesartan)

Blood thinner (warfarin, coumadin, eliquis, xarelto)

Another blood pressure medication (please specify):

Did you experience any health complications related to	Yes participating in a competitive sporting event?	No

Complications could be something like dizziness or fainting, pain, temporary disability, whether requiring professional treatment or not, or a health emergency.

Describe any health complications you experienced, including first aid or hospitalization.

Use as much detail as possible.

If you train for events, how long do you train and how

frequently? You can say something like: three times	per week for 3 months. If you do not train

specifically for events, leave this blank.

If you train for events, how long are your training

sessions? You can say something like: 2 hours. If you	do not train specifically for events, leave this

blank.

Total time spent training for athletic events:

Aside from training or participating in athletic events, how much total time in hours do you spend exercising in a typical week? You can say something like: 3 hours. If you do not regularly exercise, enter 0.

Aside from training or participating in athletic	Pilates events, what types of exercise do you do on a regular	Aerobics

basis (more than once a month)? Check all that apply:	Ballet or dance

Yoga

Exercise machines in a gym Light weights (dumbells)

Heavy weights (bench, kettlebells) Interval training

Recreational running less than 10 miles per week Recreational running 10-20 miles per week

Recreational running more than 20 miles per week Self-defense (like taekwondo, karate, boxing,

kickboxing)

Indoor cycling/spin Strength training Swimming

Something else: please specify	I do not exercise regularly

□	projectredcap.or

**Supplementary Material**

The first survey shown was distributed to the Aortic Athletes group to assess attitudes toward exercise and exercise habits. The second survey is the primary REDCap survey consisting of questions relating to competitive sports participation and event-related health outcomes. **Table 1** shows the various competitive sports and how many participants were symptomatic. **Table 2** describes the symptoms experienced by participants and the number of episodes.

**Supplementary Table 1.** Competitive Sports Participation and Event-Related Episodes

| **Event** | **Dynamic** | **Static** | **N** | **Symptomatic** | **(%)** |
| --- | --- | --- | --- | --- | --- |
| **Running** | Low | High | 41 | 4 (12) |  |
| **Cycling** | High | Low | 14 | 1 (7) |  |
| **Ironman or similar** | High | Low | 6 | 0 (0) |  |
| **Basketball** | High | Mid | 5 | 0 (0) |  |
| **Swimming** | High | Low | 5 | 0 (0) |  |
| **Walking or Hiking** | Mid | Low | 4 | 0 (0) |  |
| **Golf** | Low | Low | 3 | 1 (33) |  |
| **Football** | Mid | Mid | 3 | 0 (0) |  |
| **Soccer** | High | Low | 3 | 0 (0) |  |
| **Tennis** | High | Mid | 3 | 0 (0) |  |
| **Dancing** | High | Low | 2 | 0 (0) |  |
| **Mountaineering** | Mid | High | 1 | 1 (100) |  |
| **Jiu-Jitsu** | High | Low | 1 | 0 (0) |  |
| **Power lifting** | Low | High | 1 | 0 (0) |  |
| **Pickleball** | Mid | Low | 1 | 0 (0) |  |
| **Boxing** | High | High | 1 | 0 (0) |  |
| **Ice Hockey** | High | Mid | 2 | 0 (0) |  |
| **Wind Surfing** | Low | High | 1 | 0 (0) |  |
| **Sailing** | Low | High | 1 | 0 (0) |  |
| **Skiing** | Mid | High | 1 | 0 (0) |  |

| **Cricket** | Low | Low | 1 | 0 (0) |
| --- | --- | --- | --- | --- |
| **Riflery** | Low | Low | 1 | 0 (0) |
| **Squash** | High | Mid | 1 | 0 (0) |
| **Broomball** | High | Mid | 1 | 0 (0) |
| **Qigong, Adventure Racing** | High | Mid | 2 | 0 (0) |

### Data on dynamic and static components were derived from “Eligibility and Disqualification Recommendations for Competitive Athletes With Cardiovascular Disease: A Scientific Statement From the American Heart Association and American College of Cardiology” (doi: 10.1016/j.jacc.2015.09.032).

**Supplementary Table 2.** Event-Related Health Episodes

| **Health complication** | **Number of episodes** |
| --- | --- |
| Presyncope | 8 |
| Syncope | 2 |
| Visual disturbance | 2 |
| Arrhythmia or palpitations | 2 |
| Chest pain | 1 |
| Hospitalization | 1 |
